# mRNA-COVID19 vaccination can be considered safe and tolerable for frail patients

**DOI:** 10.1101/2022.01.18.22269351

**Authors:** Maria Teresa Lupo Stanghellini, Serena Di Cosimo, Massimo Costantini, Sara Monti, Renato Mantegazza, Alberto Mantovani, Carlo Salvarani, Pier Luigi Zinzani, Matilde Inglese, Fabio Ciceri, Giovanni Apolone, Gennaro Ciliberto, Fausto Baldanti, Aldo Morrone, Valentina Sinno, Franco Locatelli, Stefania Notari, Elena Turola, Diana Giannarelli, Nicola Silvestris, on behalf of the VAX4FRAIL Study Group

**Author notes:** Equal contribution as first author (Maria Teresa Lupo Stanghellini, Serena Di Cosimo). Corresponding author: Nicola Silvestris, Medical Oncology Department, IRCCS Istituto Tumori “Giovanni Paolo II”, DIMO – Universiuty of Bari, 70124 - Bari, Italy, Mail. An analysis from the Italian, multicentric, observational, prospective trial VAX4FRAIL study.

## Abstract

**Background:** Frail patients are considered at relevant risk of complications due to COVID-19 infection and, for this reason, are prioritized candidates for vaccination. As these patients were originally not included in the registration trials, fear related to vaccine side-effects and disease worsening was one of the reasons for vaccine hesitancy. Herein we report the safety profile of the prospective, multicenter, national VAX4FRAIL study (NCT04848493) to evaluate vaccines in a large trans-disease cohort of patients with solid or hematological malignancies, neurological and rheumatological diseases.

**Methods:** Between March 3^rd^ and September 2^nd^, 2021, 566 patients were evaluable for safety endpoint: 105 received the mRNA-1273 vaccine and 461 the BNT162b2 vaccine.

Frail patients were defined per protocol as patients under treatment with hematological malignancies (131), solid tumors (191), immune-rheumatological diseases (86), and neurological diseases (158), including multiple sclerosis and generalized myasthenia.

The impact of the vaccination on the health status of patients was assessed through a questionnaire focused on the first week after each vaccine dose.

**Results:** The most frequently reported moderate-severe adverse events were pain at the injection site (60.3% after the first dose, 55.4% after the second), fatigue (30.1% - 41.7%), bone pain (27.4% - 27.2%) and headache (11.8% - 18.9%).

Risk factors associated with the occurrence of severe symptoms after vaccine administration were identified through a multivariate logistic regression analysis: age was associated with severe fever presentation (younger patients vs. middle-aged vs. older ones), females presented a higher probability of severe pain at the injection site, fatigue, headache, and bone pain; the mRNA-1237 vaccine was associated with a higher probability of severe pain at the injection site and fever. After the first dose, patients presenting a severe symptom were at a relevant risk of recurrence of the same severe symptom after the second one.

Overall, 11 patients (1.9%) after the first dose and 7 (1.2%) after the second one required to postpone or suspend the disease-specific treatment. Finally, 2 fatal events occurred among our 566 patients. These two events were considered unrelated to the vaccine.

**Conclusions:** Our study reports that mRNA-COVID-19 vaccination is safe also in frail patients as expected side effects were manageable and had a minimum impact on patient care path.

**Importance:** Our study reports the safety analysis of the trial VAX4FRAIL confirming that mRNA-COVID-19 vaccination is safe in frail immunocompromised patients: expected side effects were manageable and had a minimum impact on patient care path.

**Objective:** To evaluate the safety of mRNA-COVID-19 vaccination in vulnerable patients.

**Design:** VAX4FRAIL is a national, multicentric, observational, prospective trial (start date March 3^rd^, 2021 – primary completion date September 2^nd^, 2021).

**Setting:** Multicenter prospective trial.

**Participants:** Frail patients were defined per protocol as patients under treatment with solid tumors (191), immune-rheumatological diseases (86), hematological malignancies (131), and neurological diseases (158), including multiple sclerosis and generalized myasthenia.

**Exposure:** Overall, 105 received the mRNA-1273 vaccine and 461 the BNT162b2 vaccine.

**Main Outcome:** The occurrence of adverse events after 1^st^ and 2^nd^ m-RNA-COVID-19 vaccination was analyzed. Adverse events were collected through a questionnaire comprising both open and closed questions.

Risk factors associated with the occurrence of severe symptoms after vaccine administration were identified through a multivariate logistic regression analysis: age was associated with severe fever presentation (younger patients vs. middle-aged vs. older ones), females presented a higher probability of severe pain at the injection site, fatigue, headache, and bone pain; the mRNA-1237 vaccine was associated with a higher probability of severe pain at the injection site and fever. Patients presenting a severe symptom after the first dose were at a relevant risk of recurrence of the same severe symptom after the second one.

Overall, 11 patients (1.9%) after the first dose and 7 (1.2%) after the second one was required to postpone or suspend their disease-specific treatment. Finally, 2 fatal events occurred among our 566 patients, and these two events were due to disease progression and considered unrelated to the vaccine.

**Conclusion and Relevance:** Our study reports that mRNA-COVID-19 vaccination is safe also in frail patients as expected side effects were manageable and had a minimum impact on patient care path.

**Study Registration:** A National, Multicentric, Observational, Prospective Study to Assess Immune Response to COVID-19 Vaccine in Frail Patients (VAX4FRAIL). NCT04848493 https://clinicaltrials.gov/ct2/show/NCT04848493

**Key Points:** *Question:* Can m-RNA-COVID19 vaccination be considered safe for frail patients?

*Findings:* In this national, multicentric, observational, prospective trial (NCT04848493) that included 566 frail patients, the occurrence of both local and systemic adverse events was manageable and did not negatively impact on the general treatment program.

*Meaning:* mRNA-COVID19 vaccination is safe among frail immunocompromised patients.

## Introduction

The currently authorized mRNA-COVID-19 vaccines – m-RNA-1237 Moderna (1) and BNT-162b2 Pfizer BioNTech (2) - have been evaluated in clinical trials that excluded in accordance with the current regulations, immunocompromised subjects, and restricted participation to healthy volunteers. Fragile patients were not considered in such pivotal studies despite being the subjects at greatest risk of COVID-19 complications and with the potential greatest advantage.

Patients diagnosed with solid or hematological malignancy or under immunosuppressive treatment due to rheumatological or neurological diseases are considered at high risk of COVID-19 complications and categorized as fragile (3-7). The intended acceptance of the COVID-19 vaccine in fragile patients confirms the positive attitudes towards vaccination, but questions arise around the safety of these vaccines in the setting of immune alterations engendered by their diseases and/or therapies (8-14). One of the most typical reasons for vaccine hesitancy has been fear related to vaccine side-effects and underlying disease worsening. An additional effort through public education campaigns specific to fragile patients to clarify vaccine’s safety, long-term effects, and potential health implications are still needed.

Over the past eight months, several groups have tried to answer questions about the efficacy and safety of COVID vaccination in different cohorts of fragile subjects. No safety concerns emerged, confirming the profile described in the series of healthy subjects (14-43) and pointing out an acceptable safety profile. VAX4FRAIL (44) study aimed at assessing immune responses to vaccination in a large trans-disease cohort of patients with hematological malignancies, solid tumors, neurological and rheumatological diseases. The study main objective is to assess prospectively the immunologic response to the COVID-19 vaccination in these specific subgroups, characterizing the kinetics of the immune response to the vaccination and its persistence over time. Longitudinal, prospective evaluation of the safety profile was part of this trans-disease study. Herein we report safety profile results as outlined in VAX4FRAIL trial.

## Methods

Safety analysis was performed among patients enrolled in the VAX4FRAIL trial between t0 and t2 according to the protocol, being t0 “time point 0” at first dose of vaccine and t2 “time point 2” the blood sampling 2-4 weeks after the second dose of vaccine (44). This is a national, multicentric observational prospective study conducted in Italy with the primary aim of assessing the immune response of COVID-19 vaccination in frail, immunocompromised patients.

Patients were considered for the current safety analysis if they met the general inclusion criteria: being ≥ 18 years of age, having received COVID-19 vaccination with mRNA vaccines (BNT-162b2 Pfizer-BioNTech or m-RNA-1237 Moderna vaccine), and having completed the health status assessment questionnaire after one of the two vaccine doses. Frail patients under evaluation were diagnosed with hematological malignancies, solid tumors, immune-rheumatological disease, and neurological disease. Detailed inclusion and exclusion criteria were previously reported (44) and disease stratification according to diagnosis and treatment of the primary disease.

The impact of the vaccination on the health status of patients was assessed through a questionnaire focused on the first week after each vaccine dose. The questionnaire was administered to the patients after the second vaccine dose (3-4 weeks after the first dose) and at the subsequent timing according to VAX4FRAIL protocol (2-4 weeks after the second dose).

The questionnaire assessed how much the patient was troubled by eight symptoms (pain or swelling at the injection site, fatigue, headache, bone pain, fever, enlarged lymph nodes, skin rash, insomnia, diarrhea, and nausea or vomiting). (See supplementary; Appendix 1) The answers were graded according to a 5-level scale (not at all, slightly, moderately, severely, and overwhelmingly). The patient was also asked to report and grade other symptoms possibly occurring after each vaccine dose and if he/she had to postpone or suspend therapies due to symptoms related to vaccination.

### Statistical methods

For this analysis, we grouped answers to the questionnaire in three levels of troubles: not at all, moderate (slightly or moderately), and severe (severely or overwhelmingly).

We estimated the probability of the occurrence of a severe symptom after the second dose of the vaccine according to the occurrence of a severe symptom after the first dose using Positive and Negative Predictive Values (PPV and NPV, respectively).

The probability of occurrence of a severe symptom after one of the two doses of the vaccine according to age, sex, main diagnosis, and type of vaccine was estimated in a multivariate logistic regression analysis adjusting for all variables.

## Results

### Patients

Between March 3^rd^ and September 2^nd^, 2021, 566 patients were enrolled in the study VAX4FRAIL and were eligible for this analysis. Among 566 patients who received the first dose of the mRNA vaccine, 105 received the mRNA-1273 and 461 the BNT162b2. Among the 566 patients who received the first dose, they also received the second dose (**Figure 1**).

**Figure 1.**
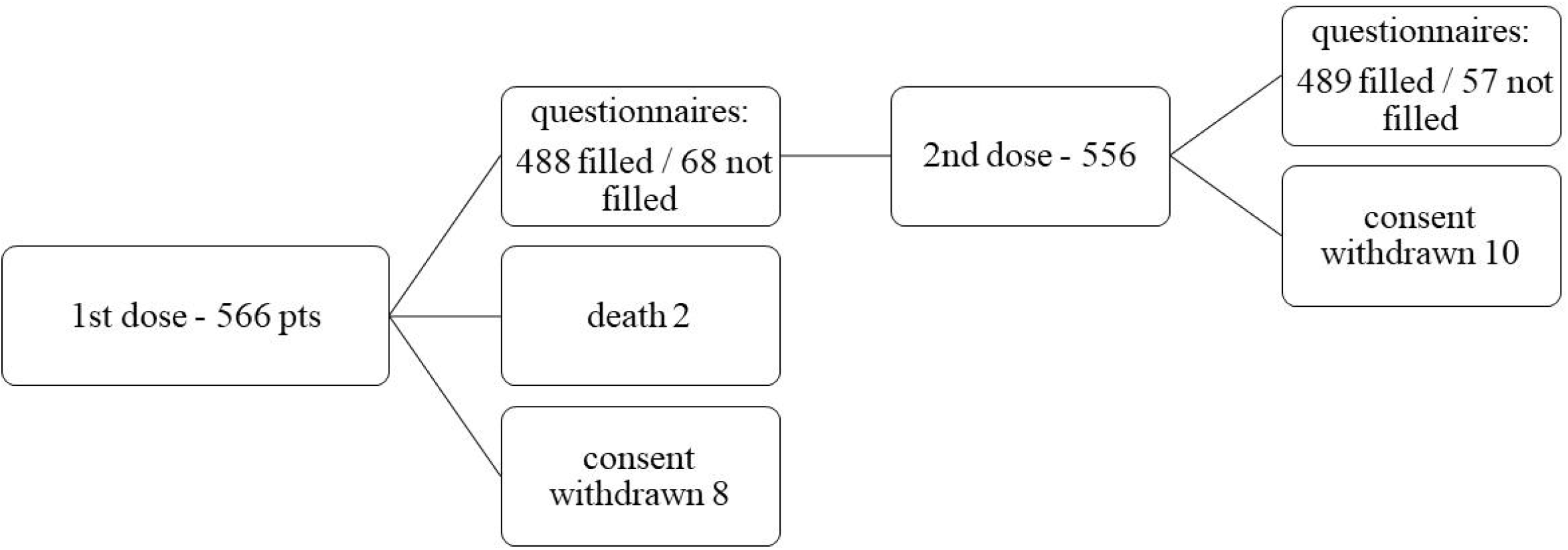
Patients’ disposition.

Overall, 488 patients after the 1^st^ dose and 489 patients after the 2^nd^ dose were evaluable for safety evaluation having complete clinical evaluation and questionnaires. Sixty-eight patients after the 1^st^ dose and 57 after the 2^nd^ dose did not fill the questionnaires. Consent withdrawn occurred in 8 cases after the 1^st^ dose and 10 cases after the 2^nd^ one.

Patient characteristics are reported in **Table 2**. Most patients (No.=299, 52.8%) were aged between 51 and 70 years, and 317 (56.0%) were females. The cohort of patients with solid tumors comprised 191 patients (33.7%), the one with neurological diseases included multiple sclerosis and generalized myasthenia 158 patients (27.9%); overall, 131 patients were diagnosed with hematological malignancies (23.1%) and 86 with immunopharmacological diseases (15.2%).

**Table 1.**
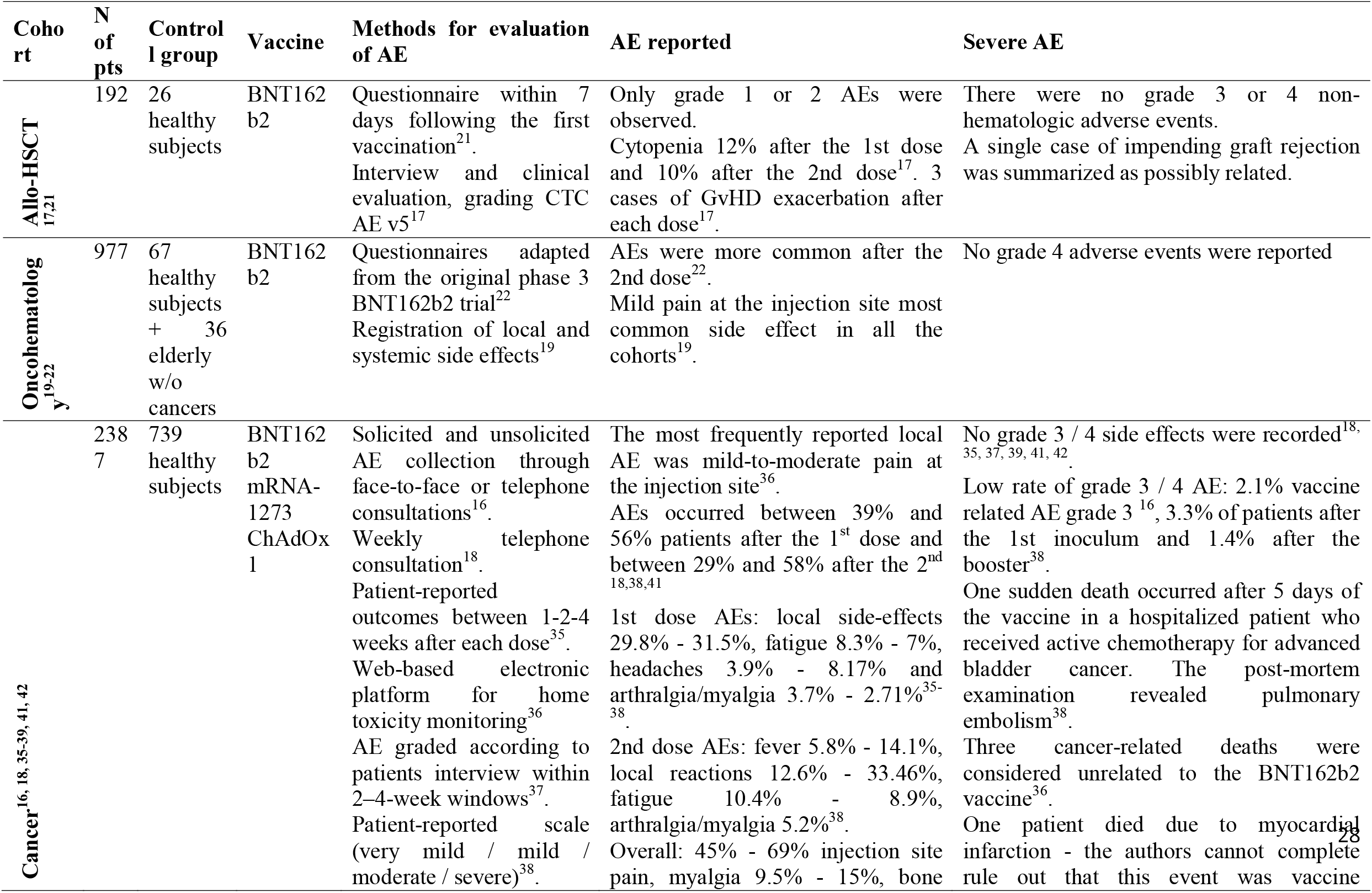
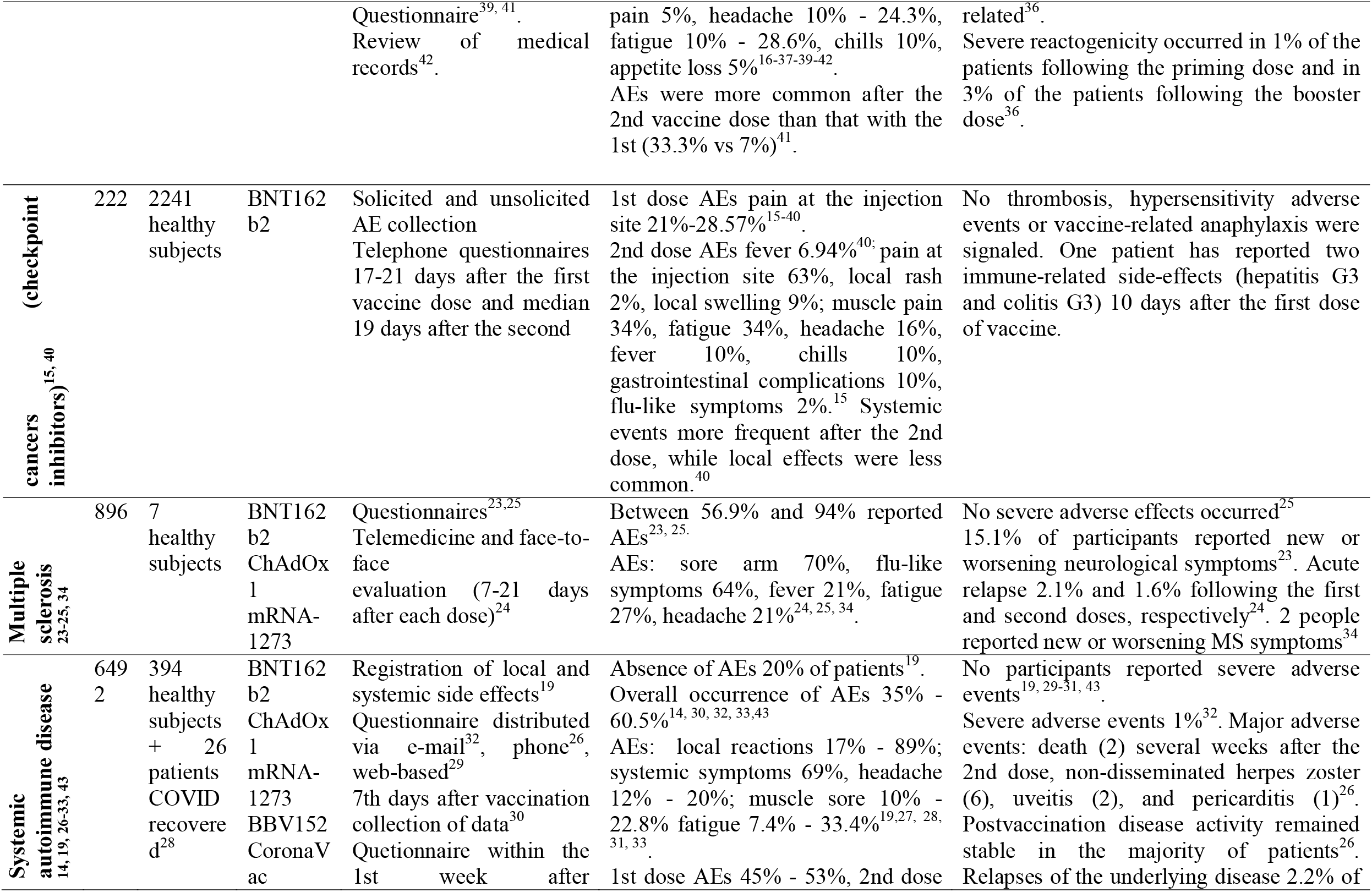
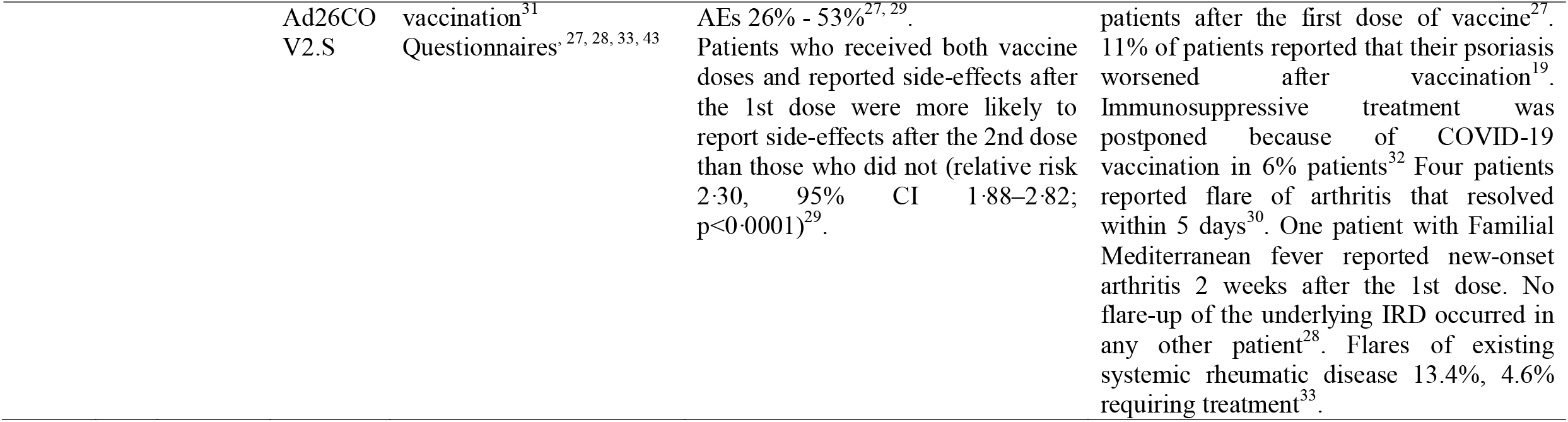
Literature review.

**Table 2.** Patients’ characteristics.

|  | N (%) |
| --- | --- |
| **Age** |  |
| 18-50 years | 160 (28.3) |
| 51-70 years | 299 (52.8) |
| >= 71 years | 107 (18.9) |
| **Patient sex** |  |
| Males | 249 (44.0) |
| Females | 317 (56.0) |
| **Main diagnosis** |  |
| Haematological malignancies | 131 (23.1) |
| Solid tumours | 191 (33.7) |
| Immunorheumatological diseases | 86 (15.2) |
| Neurological diseases | 158 (27.9) |
| **Vaccine** |  |
| mRNA-1237 Moderna | 105 (18.6) |
| BNT-162b2 Pfizer BioNTech | 461 (81.4) |
| **All patients** | 566 (100) |

### Vaccine-related adverse events

Overall, 438 (77.3%) patients reported any grade adverse events after the first dose (62, 11.0% reported as severe), and 373 (65.9%) after the second dose (87, 15.4% reported as severe). Detailed analysis of adverse events and severity is reported in **Table 3**.

**Table 3.**
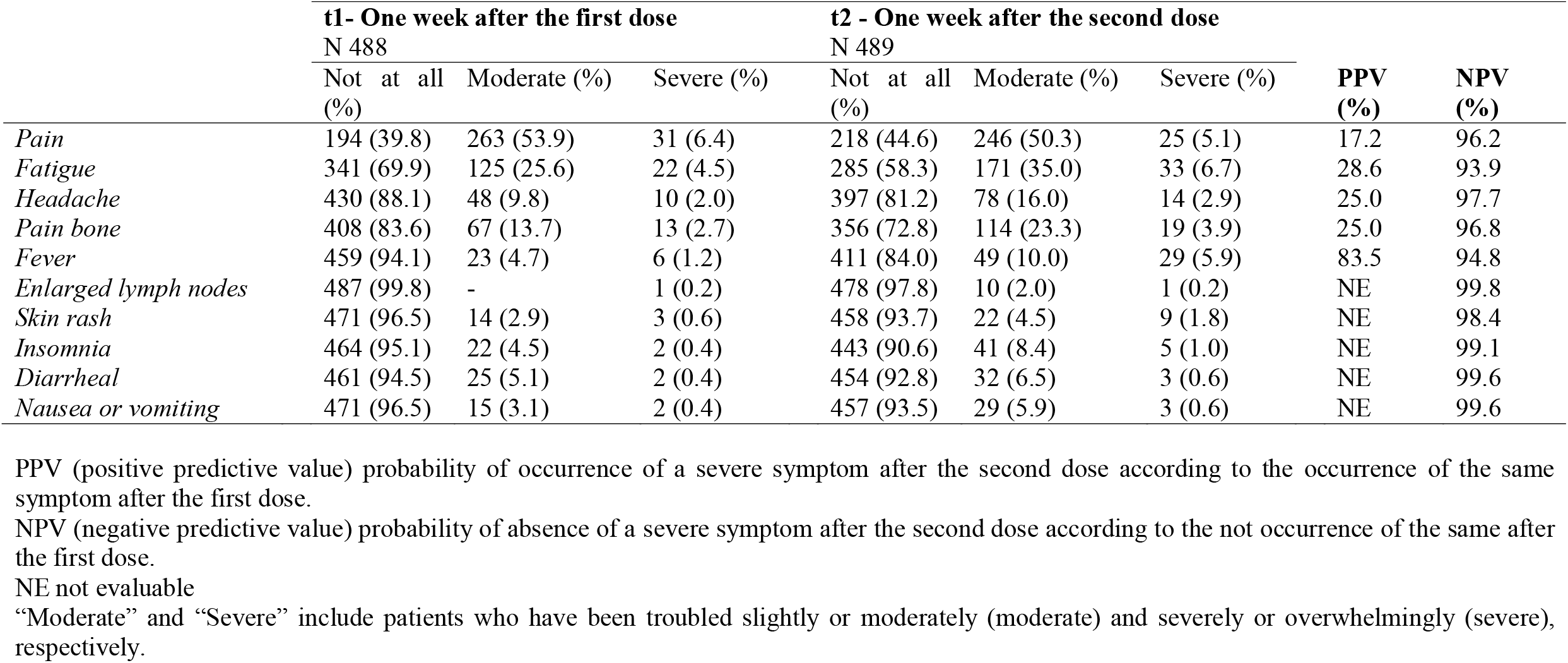
Occurrence of symptoms / adverse events over the week after vaccine administration

After the first dose (both mRNA-1273 and BNT162b2) 53.9% of patients reported moderate pain at the injection site and 6.4% severe pain: this was the most frequently reported complaint after vaccine administration. Interestingly, after the second dose - t2 - 50.3% and 5,1% reported this adverse event as moderate and severe, respectively.

At t1 25.6% of patients reported fatigue as a moderate event, 4.5% as a severe one. We observed a slight increase at t2, with 35% of patients reporting fatigue as a moderate event and 6.7% as a severe one.

Bone pain and headache were reported as moderate by 13.7% and 9.8% of patients at t1, while 2.7% and 2.0% respectively reported the symptoms as severe. After the second dose bone pain was reported as moderate by 23.3% and severe by 3.9% of patients, while headache was reported as moderate by 16% and severe by 2.9% of patients.

Only 5.9% of patients reported fever as relevant symptoms at t1 (moderate 4.7% - severe 1.2%), while at t2 the percentage of patients reporting this symptom slightly increased: 15.9% overall, with 10.0% moderate and 5.9% severe. Only a minority of patients – less than 2% - reported nausea, diarrhea, insomnia, skin rash or enlarged lymph nodes as severe manifestation both at t1 and t2.

Among unsolicited adverse events, after the first dose 5 patients reported chills (5/5 moderate), 3 itching (2 moderate, 1 severe), 2 gastro-intestinal pain (2/2 moderate), 5 myalgia (5/5 moderate), 6 dizziness (3 moderate, 3 severe), 1 drowsiness of moderate degree and 2 sweats (1 patient moderate and 1 severe). Similarly, after the second dose, 12 patients reported chills (11 moderate, 1 severe), 7 itching (5 moderate, 2 severe), 1 a moderate gastrointestinal pain, 8 myalgia (6 moderate, 2 severe), 9 dizziness (8 moderate, 1 severe), 1 drowsiness of moderate degree and 2 sweats (1 patient moderate and 1 severe). After the second dose, one patient reported metal confusion and one dysesthesia. Among the 11 patients with hematological malignancies that received an allogeneic stem cell transplantation, *graft versus host disease* (GvHD) occurrence or reactivation was not reported after the first and/or the second dose.

Of note, the absence of severe symptoms after the first dose strongly predicts the high probability of the absence of the same severe symptoms after the second dose (**Table 3**). Patients reporting severe fever after the first dose were more prone to develop the same severe symptom after the second one.

### Multivariate model for occurrence of severe symptoms

A multivariate logistic regression – adjusted by age, sex, diagnosis, and vaccine – was performed to identify risk factors associated with the occurrence of severe symptoms after vaccine administration (**Table 4**).

**Table 4.**
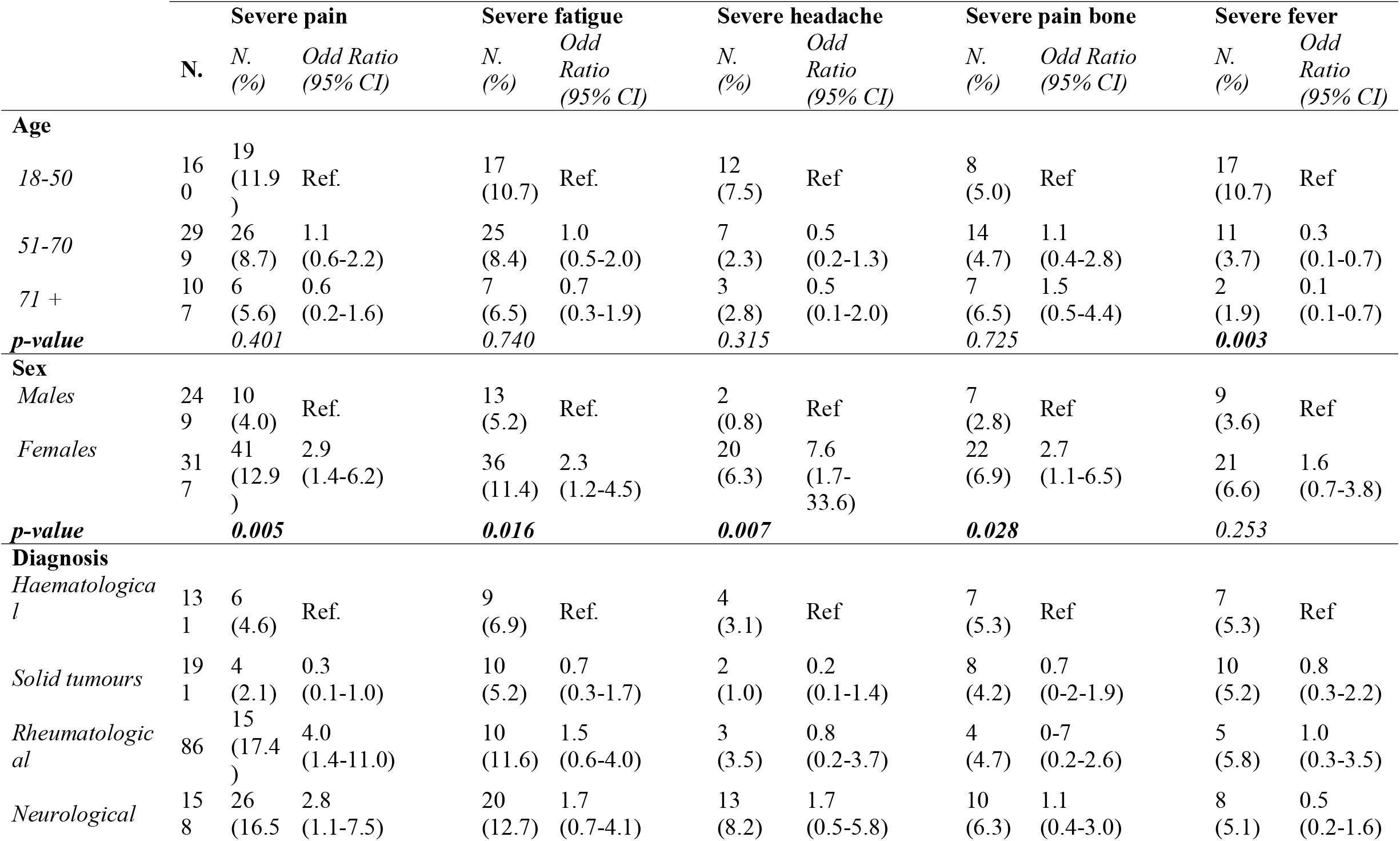
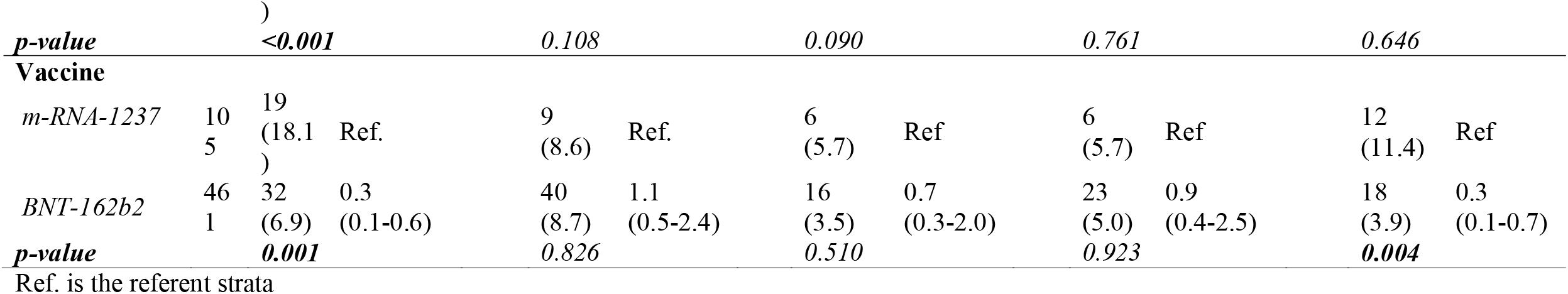
Probability of occurrence of a severe symptom after one of the two doses of vaccine estimated through a multivariate logistic regression adjusted by age, sex, diagnosis, and vaccine

Age was associated with severe fever presentation: younger patients (age 18-50 years) had a higher probability of occurrence of severe fever than middle-aged patients (age 51-70 – odd ratio 0.3, 95% confidence interval [CI] 0.1-0.7) or older ones (>/=71 – odd ratio 0.1, 95% CI 0.1-0.7) – p 0.003.

Female patients presented higher probability of severe pain at the injection site (odd ratio 2.9, 95% CI 1.4-6.2, p 0.005), severe fatigue (odd ratio 2.3, 95% CI 1.2-4.5, p 0.016), severe headache (odd ratio 7.6, 95% 1.7-33.6, p 0.007), and severe bone pain (odd ratio 2.7, 95% CI 1.1-6.5, p 0.028).

Diagnosis (hematological diseases, solid tumors, neurological diseases, rheumatological diseases) has no impact on the occurrence of severe fatigue, or headache, or bone pain or fever, but was associated with severe pain at the injection site: patients with a diagnosis of solid tumors reported severe pain less frequently than patients with hematological diseases (odds ratio 0.3, 95% CI 0.1-1.0), patients with a diagnosis of neurological or rheumatological diseases have a higher probability of occurrence of severe pain at the injection site in comparison with patients with the hematological disease (odd ration for neurological diseases 4, 95% CI 1.4-11; the odds ratio for rheumatological diseases 2.8, 95% CI 1.1-7.5) – p <0.001.

Finally, the conditional contribution made by the vaccine (mRNA-1237 versus BNT162b2) clearly showed that mRNA-1237 was associated with a higher probability of severe pain at the injection site (odds ratio 0.3, 95% CI 0.1-0.6 – p 0.001) and a higher probability of severe fever (odds ratio 0.3, 95% CI 0.1-0.7 – p 0.004).

### Impact on disease and therapy

Overall, 11 patients (1.9%) after the first dose and 7 (1.2%) after the second dose, required to postpone or suspend the disease-specific treatment due to the occurrence of symptoms associated with the vaccine. Among the 11 patients who required delays after the first dose, 7 were diagnosed with hematological disease and 4 with a solid tumor. Among the 7 patients who required delays after the second dose, 3 were diagnosed with the hematological disease and 4 with a solid tumor. No patients from the neurological diseases’ cohort or the rheumatological diseases’ cohort required treatment delay.

Fatal events, long-term sequelae, or hospitalization related to the vaccine were not registered. Two patients died from disease progression; the events were clearly reported by the investigator as not related to the vaccine administration.

## Discussion

Questionnaires based upon open and /or closed questions administered through the web-based app, phone interview, self-reporting forms, and face-to-face visits were all utilized to collect adverse events after COVID-19 vaccinations in the effort to clarify the safety of vaccines better. Absence of a univocal method to grade and collect this information make it almost impossible to directly compare the different experiences, but, of note, all the studies – both retrospective and prospective – are concordant in underlying the general perception of the absence of safety issue among distinct cohorts of frail patients (**Table 1**).

Providing information on the safety of COVID-19 vaccination in high-risk fragile populations is a duty of the international scientific community. To date, several groups are trying to answer two fundamental questions: i) if and how frail patients develop an effective response to the vaccine; ii) if the safety profile is confirmed valid in this category both with reference to toxicity and with reference to the maintenance of control of the underlying disease.

Safety profile is confirmed acceptable in all the experience reported so far in fragile patients. More systemic and local side-effects were observed after the second dose of vaccine than after the first dose. The most common local side-effects were pain at the injection site, local rash, and local swelling, whereas the most common systemic side-effects were muscle pain, fatigue, headache, fever, chills, gastrointestinal complications, and flu-like symptoms (15-16-19-20-21-23-24-25-27-28-33-34-35). Overall, the incidence of severe symptoms was low - <2.5% (16, 36, 38) – with authors reporting no severe grade 3-4 adverse events (17-18-19-21-30-35-39-40-41-42-43). In a national prospective cohort study evaluating outcomes in patients with hematological malignancies in Lithuania (22) the authors reported that adverse events were more common after the second dose, with fatigue being the most prevalent symptom (13%). No grade 4 adverse events were reported.

Among patients with multiple sclerosis, Lotan and colleagues (23) outlined how 15% of the participants reported new or worsening neurological symptoms following the vaccination, the most frequent being sensory disturbances (58.3%). Most symptoms occurred within the first 24 h after vaccination and resolved within 3 days. A total of 28 participants (77.8%) did not require any medication to treat their symptoms. Of note, no increased risk of relapse activity was noted across patients with multiple sclerosis (23-24) or autoimmune inflammatory rheumatic diseases (26-28): Boekel and coworkers clearly showed how multivariable logistic regression analyses showed similar odds for any adverse event, systemic adverse events, or moderate or severe adverse events between patients and controls, which was consistent when patients with rheumatoid arthritis or multiple sclerosis were compared with healthy controls (33).

In our experience, cohort composition was clearly defined at enrolment of patients, identifying a subset of high-risk subjects for severe COVID-19. The monitoring strategy was defined per protocol and homogeneous across the 4 categories of diseases (hematological disease, solid tumors, neurological conditions, and rheumatological diseases).

Overall, our study confirms the positive safety profile reported by other authors in both prospective and retrospective analysis (**Table 1**). Incidence of severe adverse events after vaccine administration was generally low and <3%. Most frequent complaints were pain at the injection site (severe 6.3%) and fatigue (severe 4.5%) after the first dose; pain at the injection site (severe 5.1%), fatigue (severe 6.8%), bone pain (severe 3.9%) and fever (severe 6%) after the second dose.

Patients experiencing a severe symptom after the first dose were more likely to report the same after the second one. Similarly, the absence of a severe symptom after the first dose was significantly associated with an absence of the same symptoms also after the second dose. This observation should be taken in consideration during the counseling of patients: patients should be reassured about the possibility of adopting a preventive strategy to reduce the burden of symptoms and on the not-unexpected onset of the symptom itself.

Moreover, the definition of predisposing factors to an increased possibility of presenting specific adverse events (e.g., younger patients are more likely to experience fever) can help both in the implementation of preventive measures (e.g., pre-emptive administration of painkiller drugs or antifever), in the optimization of counseling (e.g., if a patient experienced adverse events after the first shot he should be advised that the recurrence of the same after the second is very likely and that this is not unexpected, moreover strategies to counterbalance this inconvenience can be applied), and in the planning of therapies and/or vaccination itself.

Of note, less than 2% of patients required a delay or a suspension of the ongoing or planned treatment of the underlying diseases due to vaccination. This pointed out the positive safety profile of the vaccine strategy and – furthermore – this aspect should be discussed with patients to confirm the absence of impact on the whole therapy program. Whatever the underlying disease (a hematological cancer, a solid tumor, a rheumatological disease or a neurological disease) the mRNA-COVID-19 vaccine should be considered a crucial step to allow a safe program of treatment more than a possible obstacle or danger to pursue the control of the disease, being the safety profile reassuring. Fatal events or hospitalization related to the vaccine were not registered in our cohort of high-risk frail patients.

Despite the absence of safety concerns emerging from our study, we believe it is mandatory to maintain constant and careful surveillance concerning post-vaccination adverse events. This can only contribute to the reliability that the vaccination strategy pursues and represents a cornerstone of general medical practice.

As outlined by several authors (3-9), one of the most common reasons for vaccine hesitancy was fear related to vaccine side-effects and disease worsening: we can confirm that vaccine-side effects were both manageable and in line with previously reported in the general population, without the occurrence of unexpected events and no concern related to worsening of the underlying disease or the need to delay treatment.

## Conclusion

Frail patients’ candidates to mRNA-COVID-19 vaccination should be reassured about the safety profile of vaccine strategy: adverse events were in line with the report from the healthy cohort of subjects and national observatories, no evidence of worsening of the underlying disease was reported and no concern on the adherence to the treatment program of the disease itself emerged from our prospective multicenter national study. Pursuing careful and timely monitoring of expected and unexpected adverse events represents a gold standard of modern medicine and can only support evidence-based medicine.

## Data Availability

All data produced in the present work are contained in the manuscript

## Article Information

### Corresponding Author

Nicola Silvestris

### Author Contributions

MC, VS, and ET had full access to all the data in the study and took responsibility for the integrity of the data and the accuracy of the data analysis.

### Concept and design

MTLP, SDC, MC, SM, NS, GA, AM.

### Acquisition, analysis, or interpretation of data

MTLS, SDC, MC, SM, NS.

### Drafting of the manuscript

MTLS, SDC, SM, MC, NS.

### Critical revision of the manuscript for important intellectual content

All authors.

### Statistical analysis

MC, DG, ET.

### Administrative, technical, or material support

VS, ET.

### Additional Contributions

We are indebted for their precious support to this study the Research Director Dr. Giuseppe Ippolito, and the Deputy General Director for Health Research and Innovation Dr. Gaetano Guglielmi, from the Italian Ministry of Health. We would also like to thank the patients who will be enrolled in the VAX4FRAIL study and their families, and all those who will be actively involved in their continuous care, study data collection and analysis, and ultimately in the scientific production that will result from this research.

## THE VAX4FRAIL STUDY GROUP

**PRINCIPAL INVESTIGATORS** (alphabetical order): Giovanni Apolone (Fondazione IRCCS Istituto Nazionale dei Tumori di Milano); Alberto Mantovani (IRCCS Istituto Clinico Humanitas, Milano).

**SCIENTIFIC COORDINATOR**: Massimo Costantini (Fondazione IRCCS Istituto Nazionale dei Tumori di Milano).

**STEERING COMMITTEE** (alphabetical order): Chiara Agrati (IRCCS Istituto per le Malattie Infettive Lazzaro Spallanzani, Roma); Giovanni Apolone (Fondazione IRCCS Istituto Nazionale dei Tumori di Milano); Fabio Ciceri (IRCCS Ospedale San Raffaele, Milano); Gennaro Ciliberto (IRCCS Istituto Nazionale Tumori Regina Elena, Roma); Massimo Costantini (Fondazione IRCCS Istituto Nazionale dei Tumori di Milano); Franco Locatelli (Università La Sapienza, Roma); Alberto Mantovani (IRCCS Istituto Clinico Humanitas, Milano); Fausto Baldanti (Fondazione IRCCS Policlinico San Matteo di Pavia); Aldo Morrone (Istituto Dermatologico San Gallicano IRCCS, Roma); Carlo Salvarani (Azienda USL-IRCCS Reggio Emilia); Nicola Silvestris (IRCCS Istituto Tumori “Giovanni Paolo II”, Bari); Fabrizio Tagliavini (Fondazione IRCCS Istituto Neurologico Carlo Besta, Milano); Antonio Uccelli (Ospedale Policlinico San Martino IRCCS, Genova); Pier Luigi Zinzani (IRCCS Azienda Ospedaliero-Universitaria di Bologna).

## DISEASE GROUPS

1. HAEMATOLOGICAL MALIGNANCIES Referent: Paolo Corradini (Fondazione IRCCS Istituto Nazionale dei Tumori, Milano);
2. SOLID TUMORS Referent: Gennaro Ciliberto (IRCCS Istituto Nazionale Tumori Regina Elena, Roma);
3. IMMUNORHEUMATOLOGICAL DISEASES Referent: Carlo Salvarani (Azienda USL IRCCS Reggio Emilia);
4. NEUROLOGICAL DISEASES: Referent: Antonio Uccelli (Ospedale Policlinico San Martino IRCCS, Genova); Renato Mantegazza (Fondazione I.R.C.C.S Istituto Neurologico Carlo Besta (INCB), Milano).

## IMMUNOLOGICAL GROUP

### REFERENTS

Chiara Agrati (IRCCS Istituto per le Malattie Infettive Lazzaro Spallanzani, Roma); Maria Rescigno (IRCCS Istituto Clinico Humanitas, Milano); Daniela Fenoglio (Ospedale Policlinico San Martino IRCCS, Genova);

### PARTICIPANTS

Roberta Mortarini (Fondazione IRCCS Istituto Nazionale dei Tumori di Milano); Cristina Tresoldi (IRCCS Ospedale San Raffaele, Milano); Laura Conti (IRCCS Istituto Nazionale Tumori Regina Elena, Roma); Stefania Croci (Azienda USL IRCCS Reggio Emilia); Fausto Baldanti (Fondazione IRCCS Policlinico San Matteo di Pavia); Vito Garrisi (IRCCS Istituto Tumori “Giovanni Paolo II”, Bari); Fulvio Baggi (Fondazione IRCCS Istituto Neurologico Carlo Besta, Milano); Francesca Bonifazi (IRCCS Azienda Ospedaliero-Universitaria di Bologna); Fulvia Pimpinelli (Istituto Dermatologico San Gallicano IRCCS, Roma).

## INMI CENTRALIZED LABORATORY (INMI Lazzaro Spallanzani – IRCCS, Roma) (alphabetical order)

Enrico Girardi (Scientific Director), Aurora Bettini; Veronica Bordoni; Concetta Castilletti; Eleonora Cimini; Rita Casetti; Francesca Colavita; Flavia Cristofanelli; Massimo Francalancia; Simona Gili; Giulia Gramigna; Germana Grassi; Daniele Lapa; Sara Leone; Davide Mariotti; Giulia Matusali; Silvia Meschi; Stefania Notari; Enzo Puro; Marika Rubino; Alessandra Sacchi; Eleonora Tartaglia

## CLINICAL TASK FORCE

Paolo Corradini, Silvia Damian, Filippo de Braud (Fondazione IRCCS Istituto Nazionale dei Tumori di Milano); Maria Teresa Lupo Stanghellini, Lorenzo Dagna, Francesca Ogliari, Massimo Filippi (IRCCS Ospedale San Raffaele: Milano); Giulia Piaggio (IRCCS Istituto Nazionale Tumori Regina Elena, Roma); Elena Azzolini, Chiara Pozzi, Luca Germagnoli, Carlo Selmi, Maria De Santis, Carmelo Carlo-Stella, Alexia Bertuzzi, Francesca Motta, Angela Ceribelli (IRCCS Istituto Clinico Humanitas, Milano);

**Fondazione IRCCS Policlinico San Matteo di Pavia:** Sara Monti,

**Istituto Dermatologico San Gallicano IRCCS, Roma**;

**Azienda USL-IRCCS Reggio Emilia:** Maria Grazia Catanoso,

Rosa Divella, Antonio Tufaro, Vito Garrisi, Sabina Delcuratolo, Mariana Miano (IRCCS Istituto Tumori “Giovanni Paolo II”, Bari;

**Fondazione IRCCS Istituto Neurologico Carlo Besta, Milano**;

**Ospedale Policlinico San Martino IRCCS, Genova**;

Marco Fusconi, Vittorio Stefoni, Maria Abbondanza Pantaleo (IRCCS Azienda Ospedaliero-Universitaria di Bologna).

## STATISTICAL COMMITTEE

Diana Giannarelli (IRCCS Istituto Nazionale Tumori Regina Elena, Roma).

## e-CRF AND MONITORING REFERENT

Valentina Sinno, Serena Di Cosimo (Fondazione IRCCS Istituto Nazionale dei Tumori di Milano).

## PROJECT MANAGERS OF THE STUDY

### REFERENTS

Elena Turola, Azienda USL-IRCCS di Reggio Emilia.

### PARTICIPANTS

Iolanda Pulice, Roberta Mennitto Fondazione IRCCS Istituto Nazionale dei Tumori, Milano); Stefania Trinca (IRCCS Ospedale San Raffaele, Milano); Giulia Piaggio (IRCCS Istituto Nazionale Tumori Regina Elena, Roma); Chiara Pozzi (IRCCS Istituto Clinico Humanitas, Milano);

Irene Cassaniti (Fondazione IRCCS Policlinico San Matteo, Pavia); Alessandro Barberini (Istituto Dermatologico San Gallicano IRCCS, Roma); Arianna Belvedere (Azienda USL-IRCCS Reggio Emilia);

Sabina Del Curatolo (IRCCS Istituto Tumori “Giovanni Paolo II”, Bari); Rinaldi Elena, Federica Bortone (Fondazione IRCCS Istituto Neurologico Carlo Besta, Milano); Maria Giovanna Dal Bello (Ospedale Policlinico San Martino IRCCS, Genova); Silvia Corazza (IRCCS Azienda Ospedaliero-Universitaria, Bologna).

### Institutional Review Boards

The study, according to the National COVID-19 procedures, was approved by the Italian Regulatory Agency (AIFA) and by the Ethics Committee of IRCCS L. Spallanzani (code 304,2021).

