## supplemental for "mRNA-COVID19 vaccination can be considered safe and tolerable for frail patients"

**Appendix 1:** The questionnaire

This form must be filled in:

- at T1 (after the second dose of vaccine)

- at T2 (after the blood sampling 5-8 weeks after the first dose of vaccine)

The questionnaire is self-administered and is referred to the experience of the patient in the week after the vaccine administration

Over the week after vaccine administration, have you been affected by these symptoms?

1. pain or swelling at the injection site:

|  | 0 |  | Not at all |
| --- | --- | --- | --- |
|  | 1 |  | Slightly |
|  | 2 |  | Moderately |
|  | 3 |  | Severely |
|  | 4 |  | Overwhelming |

2. Fatigue

|  | 0 |  | Not at all |
| --- | --- | --- | --- |
|  | 1 |  | Slightly |
|  | 2 |  | Moderately |
|  | 3 |  | Severely |
|  | 4 |  | Overwhelming |

3. Headache:

|  | 0 |  | Not at all |
| --- | --- | --- | --- |
|  | 1 |  | Slightly |
|  | 2 |  | Moderately |
|  | 3 |  | Severely |
|  | 4 |  | Overwhelming |

4. Pain or muscles bone:

|  | 0 |  | Not at all |
| --- | --- | --- | --- |
|  | 1 |  | Slightly |
|  | 2 |  | Moderately |
|  | 3 |  | Severely |
|  | 4 |  | Overwhelming |

5. Fever:

|  | 0 |  | Not at all |
| --- | --- | --- | --- |
|  | 1 |  | Slightly |
|  | 2 |  | Moderately |
|  | 3 |  | Severely |
|  | 4 |  | Overwhelming |

6. Enlarged lymph nodes:

|  | 0 |  | Not at all |
| --- | --- | --- | --- |
|  | 1 |  | Slightly |
|  | 2 |  | Moderately |
|  | 3 |  | Severely |
|  | 4 |  | Overwhelming |

7. Skin rash:

|  | 0 |  | Not at all |
| --- | --- | --- | --- |
|  | 1 |  | Slightly |
|  | 2 |  | Moderately |
|  | 3 |  | Severely |
|  | 4 |  | Overwhelming |

8. Insomnia:

|  | 0 |  | Not at all |
| --- | --- | --- | --- |
|  | 1 |  | Slightly |
|  | 2 |  | Moderately |
|  | 3 |  | Severely |
|  | 4 |  | Overwhelming |

9. Diarrheal:

|  | 0 |  | Not at all |
| --- | --- | --- | --- |
|  | 1 |  | Slightly |
|  | 2 |  | Moderately |
|  | 3 |  | Severely |
|  | 4 |  | Overwhelming |

10. Nausea e/o vomiting:

|  | 0 |  | Not at all |
| --- | --- | --- | --- |
|  | 1 |  | Slightly |
|  | 2 |  | Moderately |
|  | 3 |  | Severely |
|  | 4 |  | Overwhelming |

11. Other (specify):

|  | 0 |  | Not at all |
| --- | --- | --- | --- |
|  | 1 |  | Slightly |
|  | 2 |  | Moderately |
|  | 3 |  | Severely |
|  | 4 |  | Overwhelming |

Did the patient had to postpone or suspend therapies due to symptoms related to vaccination?

□ yes	□ no
